# Comparative Performance of Clinical Scoring Systems for Early Mortality Prediction in Blunt Traumatic Brain Injury

**DOI:** 10.64898/2026.07.03.26357217

**Authors:** Mostafa Abdollahi Sarvi

## Abstract

**Background:** Early risk stratification in traumatic brain injury (TBI) is essential for timely triage, resource allocation, and clinical decision-making within the critical first hours of admission. This study aimed to compare the predictive performance of five established clinical scoring systems includes the Glasgow Coma Scale (GCS), Revised Trauma Score (RTS), Mechanism, GCS, Age, and Arterial Pressure (MGAP) score, Modified Early Warning Score (MEWS), and Rapid Emergency Medicine Score (REMS) for early mortality prediction in patients with blunt TBI.

**Methods:** This single-center retrospective observational cohort study evaluated 444 patients aged 18 to 89 years with blunt TBI admitted to the intensive care unit of a tertiary trauma center in Tehran, Iran, between March 2022 and March 2025. The primary outcome was early mortality, defined as death within 24 hours of admission. Discriminative performance was assessed using the area under the receiver operating characteristic curve (AUC) with 95% confidence intervals (CI) derived via bootstrap resampling (1,000 iterations). Pairwise comparisons of AUCs were conducted, and optimal diagnostic cutoffs were identified using the Youden Index.

**Results:** Within 24 hours of hospital admission, early mortality occurred in 97 patients (21.8%), while 347 patients (78.2%) survived. The trauma-specific and neurological scoring systems demonstrated the highest discriminative capacities: RTS achieved the highest accuracy (AUC **=** 0.676, 95% CI: 0.617–0.737), followed closely by GCS (AUC = 0.669, 95% CI: 0.608–0.727) and MGAP (AUC = 0.657, 95% CI: 0.594–0.724). General physiological scores exhibited lower performance, with MEWS achieving an AUC of 0.651 (95% CI: 0.595–0.707) and REMS demonstrating the lowest discriminative ability (AUC = 0.601, 95% CI: 0.538–0.659). Pairwise analysis confirmed that RTS GCS and MGAP significantly outperformed REMS, though no statistically significant differences were observed among RTS, GCS, and MGAP themselves. All evaluated systems demonstrated only modest overall predictive performance (AUC < 0.70).

**Conclusion:** Trauma-specific and neurologically oriented scoring systems (RTS, GCS, and MGAP) provide superior and comparable prognostic accuracy for 24-hour mortality in blunt TBI compared to general emergency scores like REMS. However, the absolute predictive power of all evaluated models remains modest. Traditional systems relying on static admission variables fail to capture the dynamic, multifactorial nature of secondary brain injury, highlighting the critical need for multidimensional prognostic tools incorporating physiological time-series data or machine learning algorithms.

## 1. Introduction

Traumatic brain injury (TBI) remains a major public health concern and is one of the leading causes of mortality and long-term disability worldwide [1, 2]. The burden of TBI is particularly pronounced among young adults and economically active populations, resulting in substantial healthcare expenditures and societal costs [1]. Despite advances in prehospital care, neurocritical care, and neurosurgical management, early mortality following TBI continues to represent a significant clinical challenge, particularly in patients presenting with severe injury and physiological instability [3, 4].

Early identification of patients at increased risk of mortality is essential for timely clinical decision-making, triage, resource allocation, and prognostic counseling [5, 6]. Various clinical scoring systems have been developed to facilitate rapid assessment of injury severity and mortality risk in emergency and trauma settings [6, 7]. Among these, the Glasgow Coma Scale (GCS) remains the most widely utilized neurological assessment tool and serves as a cornerstone in the initial evaluation of patients with TBI [8]. However, neurological assessment alone may not fully reflect the complex physiological responses and hemodynamic alterations that accompany traumatic injury and influence patient outcomes [9].

To address this limitation, several composite scoring systems incorporating demographic and physiological variables have been developed to facilitate rapid risk stratification in trauma and emergency settings [5]. The Revised Trauma Score (RTS), originally introduced as a physiologically based trauma assessment tool, integrates the Glasgow Coma Scale, systolic blood pressure, and respiratory rate to estimate injury severity and predict trauma outcomes [6]. Building upon the prognostic importance of age and injury mechanism, the Mechanism, Glasgow Coma Scale, Age, and Arterial Pressure (MGAP) score was subsequently developed as a simplified prehospital and emergency department scoring system for early mortality prediction in trauma patients [6]. Both RTS and MGAP were specifically designed for trauma populations and have been validated across diverse injury settings [5, 6].

In contrast, the Modified Early Warning Score (MEWS) and the Rapid Emergency Medicine Score (REMS) originated from efforts to identify critically ill patients within general emergency and acute care populations. MEWS utilizes routinely available physiological parameters, including heart rate, respiratory rate, systolic blood pressure, temperature, and level of consciousness, to detect clinical deterioration and guide escalation of care. REMS was developed as a simplified alternative to more complex critical care severity indices and incorporates age, mean arterial pressure, heart rate, respiratory rate, oxygen saturation, and neurological status to predict in-hospital mortality. Although MEWS and REMS have demonstrated prognostic value in heterogeneous emergency department cohorts, their applicability and comparative performance in patients with blunt traumatic brain injury remain less clearly defined [9].

On the other hand, these scoring systems have demonstrated prognostic utility in diverse trauma and emergency care populations, their comparative performance in patients with blunt traumatic brain injury remains incompletely understood [6]. Furthermore, direct head-to-head comparisons among neurological, trauma-specific, and general emergency scoring systems within a single blunt TBI cohort are limited. Therefore, the present study aimed to compare the predictive performance of GCS, MGAP, RTS, MEWS, and REMS for early mortality prediction in patients with blunt traumatic brain injury. We hypothesized that trauma-specific and neurological scoring systems would demonstrate superior discriminative ability compared with general emergency scoring systems for predicting mortality within the first 24 hours following hospital admission.

## 2. Method

### 2.1 Study Design and Setting

This study was designed as a single-center retrospective observational cohort study conducted among patients with blunt traumatic brain injury (TBI) admitted to the intensive care unit (ICU) of a tertiary referral trauma center in Tehran Province, Iran, between March 2022 and March of 2025.

### 2.2 Study Population

The study population included patients aged 18 to 89 years who were admitted to the ICU following blunt traumatic brain injury and received standard diagnostic and therapeutic management. Only patients with complete documentation of required clinical and physiological variables at the time of admission were eligible for inclusion. All injuries were blunt in nature.

A total of 450 patients were initially identified. After exclusion of records with incomplete or missing essential variables required for score calculation, 444 patients were included in the final analytical cohort.

### 2.3 Data Collection

Demographic, clinical, and physiological data were extracted from electronic and paper-based medical records using a standardized data extraction form. Variables included age, sex, Glasgow Coma Scale (GCS), respiratory rate, peripheral oxygen saturation (SpO_2_), systolic blood pressure (SBP), diastolic blood pressure (DBP), mean arterial pressure (MAP), heart rate (HR), body temperature, and level of consciousness assessed using the AVPU (Alert, Voice, Pain, Unresponsive) scale. The primary outcome was early mortality, defined as death occurring within 24 hours of hospital admission. Patients surviving beyond 24 hours were classified as survivors in our scope.

### 2.4 Clinical Scoring Systems

Five established clinical scoring systems were evaluated for their ability to predict early mortality:

- **Glasgow Coma Scale (GCS):** A widely used neurological scale ranging from 3 to 15 points based on eye, verbal, and motor responses [10].
- **Mechanism, Glasgow Coma Scale, Age, and Arterial Pressure (MGAP):** A trauma specific prognostic score incorporating injury mechanism, GCS, age, and systolic blood pressure [11].
- **Revised Trauma Score (RTS):** A physiologically based trauma scoring system derived from coded values of GCS, systolic blood pressure, and respiratory rate [12].
- **Modified Early Warning Score (MEWS):** A general physiological deterioration score based on systolic blood pressure, heart rate, respiratory rate, temperature, and level of consciousness [13].
- **Rapid Emergency Medicine Score (REMS):** A prognostic scoring system incorporating age, mean arterial pressure, heart rate, respiratory rate, oxygen saturation, and neurological status [14].

All scores were calculated according to their original published definitions using admission data [10-14].

### 2.5 Statistical Analysis

Continuous variables were reported as means with standard deviations or medians with interquartile ranges, depending on distribution. Categorical variables were presented as frequencies and percentages.

The discriminative performance of each scoring system for predicting 24-hour mortality was assessed using receiver operating characteristic (ROC) curve analysis. Area under the curve (AUC) values were calculated with 95% confidence intervals obtained via bootstrap resampling with 1,000 iterations.

Pairwise comparisons of AUCs were performed using non-parametric bootstrap methods to assess statistically significant differences between scoring systems. Optimal cutoff values were determined using the Youden Index, defined as sensitivity + specificity − 1 [15]. Sensitivity and specificity were calculated for each score at its optimal threshold.

All analyses were conducted using Python (Python Software Foundation, Wilmington, DE, USA) with pandas, NumPy, scikit-learn [16], and matplotlib libraries. A two-sided p-value < 0.05 was considered statistically significant.

### 2.6 Ethical Considerations

This study was conducted in accordance with the Declaration of Helsinki and was approved by the Ethics Committee of Iran University of Medical Sciences. Given the retrospective and non-interventional nature of the study, informed consent was waived. All data were de-identified prior to analysis to ensure patient confidentiality. Data handling and storage complied with institutional and national regulations on medical data privacy and security.

## 3. Results

### 3.1 Baseline Characteristics

A total of 444 patients with blunt traumatic brain injury were included in the final analysis. The cohort consisted of patients aged 18 to 89 years admitted to the intensive care unit of a tertiary referral trauma center in Tehran Province, Iran. Among these patients, 97 (21.8%) experienced early mortality within 24 hours of admission, while 347 (78.2%) survived beyond 24 hours. Table 1 summarizes the baseline demographic, physiological, and neurological characteristics of the study population stratified by 24-hour mortality. Significant differences were observed between survivors and non-survivors in GCS, systolic blood pressure, mean arterial pressure, heart rate, and oxygen saturation (all p < 0.05). In contrast, age, diastolic blood pressure, respiratory rate, and temperature did not show statistically significant differences between groups. Male sex distribution was higher in both groups but did not differ significantly (p = 0.346). AVPU level demonstrated a significant association with mortality status (p < 0.001), with a higher proportion of lower consciousness states observed in the non-survivor group.

**Table 1.**
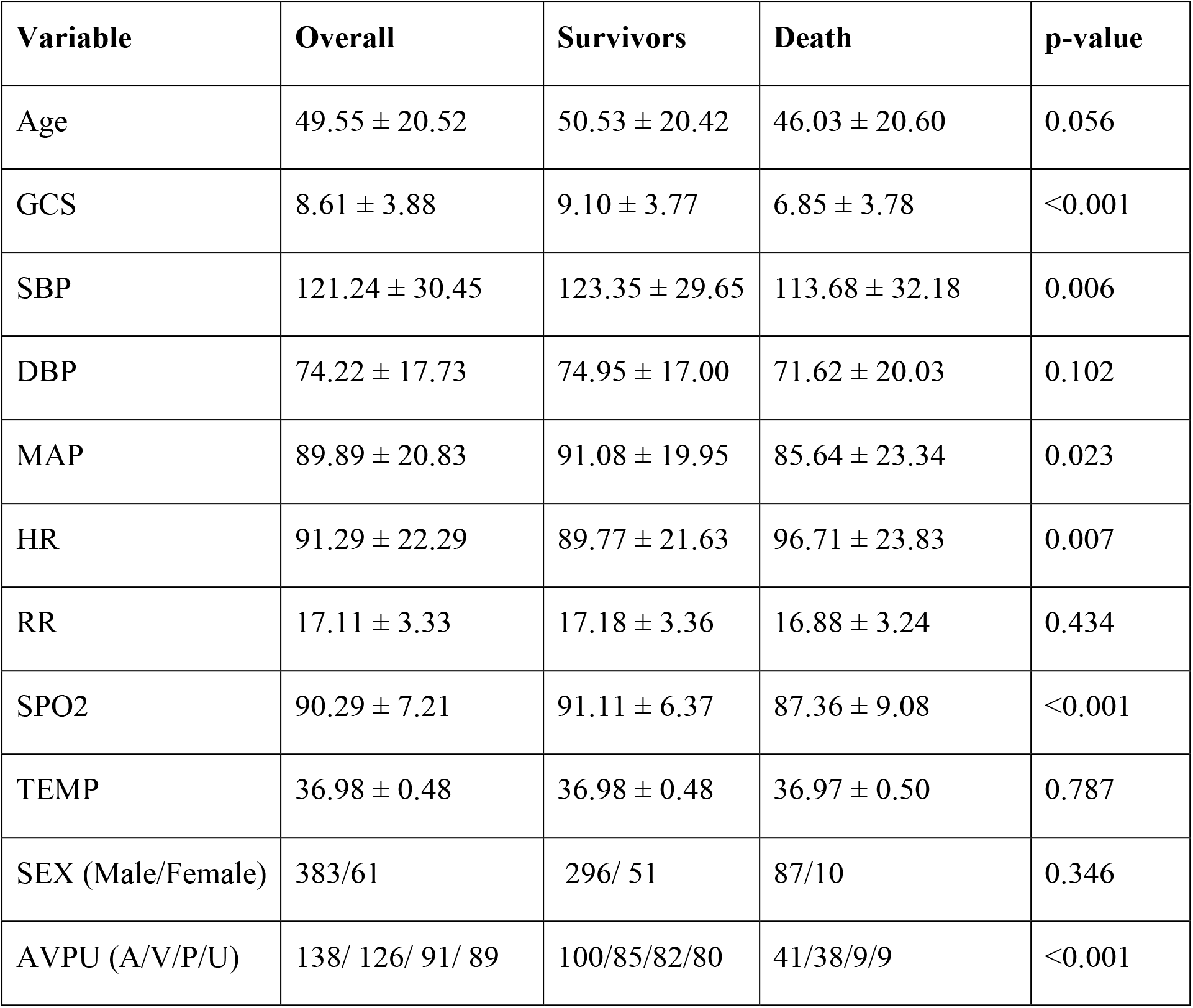
Baseline and comparison of demographic and clinical characteristics.

### 3.2 Predictive Performance of Clinical Scoring Systems

The characteristics of clinical scoring systems among survivors and non-survivors is presented in Table 2. Significant differences were observed between outcome groups for all evaluated scoring systems (all *p* < 0.001). Patients who died within 24 hours had lower mean GCS, MGAP, and RTS values, whereas higher REMS and MEWS values were observed among non-survivors.

**Table 2.** Comparison of Clinical Scoring Systems Between Survivors and Non-Survivors. Values are presented as mean ± standard deviation. P-values were calculated by comparing survivors and non-survivors. Lower GCS, MGAP, and RTS scores and higher REMS and MEWS scores were associated with increased 24-hour mortality.

| Score | Overall | Survivors | Death | p-value |
| --- | --- | --- | --- | --- |
| GCS | 8.61 ± 3.88 | 9.10 ± 3.77 | 6.85 ± 3.78 | <0.001 |
| MGAP | 19.80 ± 4.18 | 20.28 ± 4.04 | 18.08 ± 4.24 | <0.001 |
| RTS | 6.13 ± 1.39 | 6.32 ± 1.33 | 5.46 ± 1.37 | <0.001 |
| REMS | 6.62 ± 3.43 | 6.35 ± 3.36 | 7.57 ± 3.54 | <0.001 |
| MEWS | 3.57 ± 2.17 | 3.33 ± 2.12 | 4.44 ± 2.14 | <0.001 |

The discriminative performance of the evaluated scoring systems for predicting 24-hour mortality is summarized in Table 3 and illustrated as AUC-ROC in Figure 1. Among the assessed scores, the Revised Trauma Score (RTS) demonstrated the highest predictive accuracy, with an AUC of 0.676 (95% CI: 0.617–0.737), followed closely by the Glasgow Coma Scale (GCS) (AUC = 0.669, 95% CI: 0.608–0.727) and the Mechanism, Glasgow Coma Scale, Age, and Arterial Pressure (MGAP) score (AUC = 0.657, 95% CI: 0.594–0.724). The Modified Early Warning Score (MEWS) demonstrated similar but slightly lower discrimination (AUC = 0.651, 95% CI: 0.595–0.707), whereas the Rapid Emergency Medicine Score (REMS) showed the lowest predictive performance (AUC = 0.601, 95% CI: 0.538–0.659).

**Table 3.** Discriminative Performance of Clinical Scoring Systems for Prediction of 24-Hour Mortality. Area under the receiver operating characteristic curve (AUC) and corresponding 95% confidence intervals for the evaluated clinical scoring systems. Higher AUC values indicate better discrimination between survivors and non-survivors.

| Score | AUC | 95% CI |
| --- | --- | --- |
| RTS | 0.676 | 0.617–0.737 |
| GCS | 0.669 | 0.608–0.727 |
| MGAP | 0.657 | 0.594–0.724 |
| MEWS | 0.651 | 0.595–0.707 |
| REMS | 0.601 | 0.538–0.659 |

**Figure 1.**
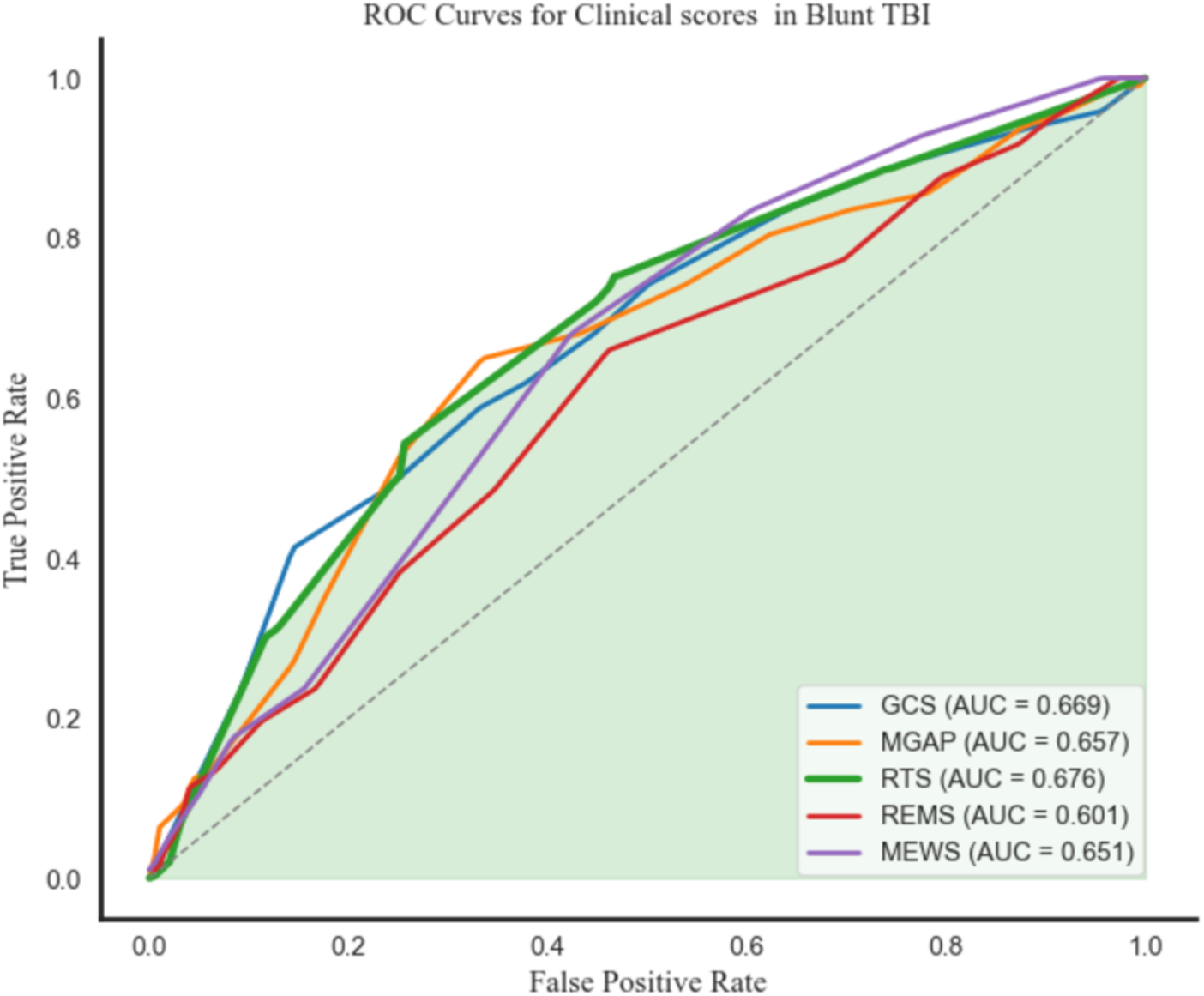
ROC Curves of Clinical Scoring Systems for Prediction of 24-Hour Mortality in Patients with Blunt TBI. ROC curves demonstrating the predictive performance of GCS, MGAP, RTS, REMS, and MEWS for 24-hour mortality. RTS achieved the highest discriminative performance (AUC = 0.676), followed by GCS (AUC = 0.669) and MGAP (AUC = 0.657). The diagonal dashed line represents the line of no discrimination (AUC = 0.50).

Overall, trauma-specific scoring systems (RTS and MGAP) and the neurological assessment represented by GCS demonstrated superior discrimination compared with general physiological scoring systems, particularly REMS. However, all evaluated scores exhibited only modest predictive performance, with AUC values below 0.70.

### 3.3 Pairwise Comparison of Predictive Performance

Pairwise comparisons of receiver operating characteristic (ROC) curves are presented in Table 4. No statistically significant differences were observed among the trauma-oriented scoring systems RTS, GCS, and MGAP (all *p* > 0.05), indicating comparable discriminative performance for predicting 24-hour mortality in patients with blunt traumatic brain injury.

Compared with REMS, RTS demonstrated significantly superior predictive performance (AUC difference = 0.076, 95% CI: 0.013–0.136, *p* = 0.012). Similarly, GCS (AUC difference = 0.070, 95% CI: 0.003–0.134, *p* = 0.038) and MGAP (AUC difference = 0.057, 95% CI: 0.003–0.112, *p* = 0.036)significantly outperformed REMS. Although MEWS demonstrated a numerically higher AUC than REMS, this difference did not reach statistical significance (*p* = 0.109).

No statistically significant differences were identified between MEWS and RTS, GCS, or MGAP (all *p* > 0.05), suggesting that despite slightly lower discrimination, MEWS performed comparably to the trauma-specific scoring systems in this cohort.

Overall, these findings indicate that RTS, GCS, and MGAP provide similar predictive accuracy, whereas REMS demonstrates significantly inferior performance compared with several trauma-focused scoring systems.

| Score 1 | Score 2 | AUC Difference | 95% CI | p-value |
| --- | --- | --- | --- | --- |
| RTS | GCS | 0.006 | -0.014 to 0.028 | 0.552 |
| RTS | MGAP | 0.020 | -0.017 to 0.056 | 0.289 |
| RTS | REMS | 0.077 | 0.013 to 0.136 | 0.014 |
| RTS | MEWS | 0.026 | -0.024 to 0.077 | 0.308 |
| GCS | MGAP | 0.013 | -0.022 to 0.048 | 0.486 |
| GCS | REMS | 0.069 | 0.003 to 0.135 | 0.043 |
| GCS | MEWS | 0.018 | -0.041 to 0.076 | 0.527 |
| MGAP | REMS | 0.057 | 0.003 to 0.112 | 0.036 |
| MGAP | MEWS | 0.005 | -0.058 to 0.067 | 0.874 |
| REMS | MEWS | -0.050 | -0.112 to 0.014 | 0.127 |

### 3.4 Sensitivity and Specificity Analysis

Optimal cutoff values derived using the Youden Index are presented in Table 5. Among the evaluated scoring systems, the Mechanism, Glasgow Coma Scale, Age, and Arterial Pressure (MGAP) score demonstrated the most balanced diagnostic performance, with relatively comparable sensitivity and specificity for predicting 24-hour mortality in patients with blunt traumatic brain injury.

**Table 5.**
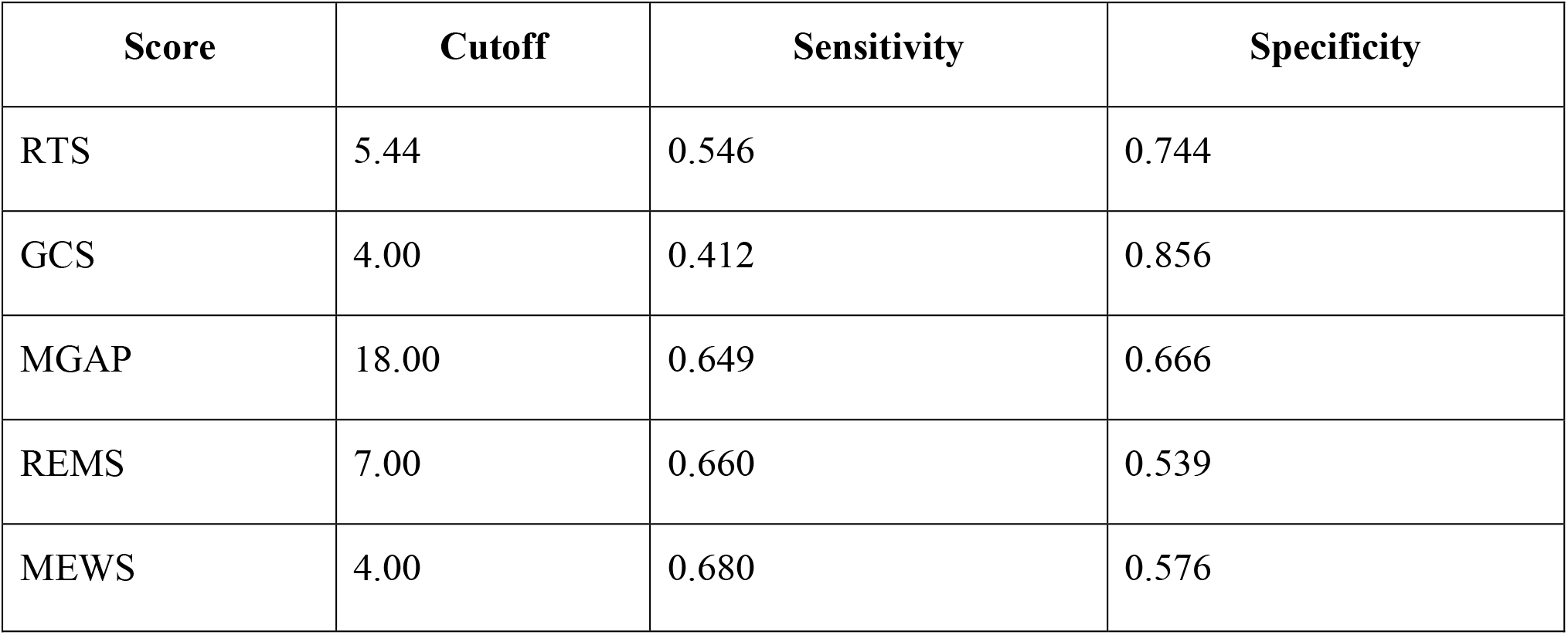
Optimal Cutoff Values and Diagnostic Performance of Clinical Scoring Systems for Prediction of 24-Hour Mortality. Higher sensitivity indicates better detection of mortality cases, whereas higher specificity indicates better identification of survivors.

| Score | Cutoff | Sensitivity | Specificity |
| --- | --- | --- | --- |
| RTS | 5.44 | 0.546 | 0.744 |
| GCS | 4.00 | 0.412 | 0.856 |
| MGAP | 18.00 | 0.649 | 0.666 |
| REMS | 7.00 | 0.660 | 0.539 |
| MEWS | 4.00 | 0.680 | 0.576 |

In contrast, the Glasgow Coma Scale (GCS) exhibited higher specificity but lower sensitivity, indicating stronger performance in correctly identifying survivors at the expense of reduced detection of early mortality cases. The Modified Early Warning Score (MEWS) demonstrated higher sensitivity but lower specificity, suggesting improved identification of high-risk patients but with a greater rate of false-positive classification.

The Rapid Emergency Medicine Score (REMS) showed moderate sensitivity but comparatively lower specificity, indicating limited discriminative balance. The Revised Trauma Score (RTS) demonstrated intermediate diagnostic performance with relatively balanced sensitivity and specificity, consistent with its overall AUC performance.

Overall, trauma-specific and neurological scoring systems (RTS, GCS, and MGAP) demonstrated superior and comparable diagnostic performance compared with general emergency scoring systems (MEWS and REMS). Among the latter, REMS consistently showed the lowest overall predictive accuracy across all evaluated analyses, including ROC-based discrimination and optimal cutoff performance.

## 4. Discussion

Traumatic brain injury (TBI) remains a leading cause of early mortality in trauma care, where timely risk stratification is essential for guiding clinical decision-making, especially in the first hours after admission [1, 2]. In this retrospective cohort of 444 patients with blunt TBI admitted to a tertiary trauma center, we evaluated the prognostic performance of commonly used clinical scoring systems and their ability to predict early mortality within 24 hours.

Early mortality in TBI is fundamentally driven by the interplay between neurological injury severity and systemic physiological failure [1]. Reduced consciousness at presentation reflects the extent of primary brain injury and disruption of cortical and brainstem function, while accompanying hemodynamic and oxygenation abnormalities reflect systemic decompensation and impaired compensatory capacity [1, 5]. Together, these findings reinforce the concept that early outcomes in TBI are not determined solely by focal neurological damage but by a global failure of integrated cerebral and systemic physiology [1].

Within this framework, trauma-specific and neurologically oriented scoring systems demonstrated better prognostic performance than general emergency scoring tools [6]. This is likely explained by their inclusion of key determinants of early TBI outcome, particularly consciousness level and cardiovascular status, which are central to the pathophysiology of secondary brain injury [1, 6]. In contrast, general emergency scoring systems were developed for heterogeneous populations and may not adequately capture the specific physiological signatures of isolated neurotrauma, limiting their discriminative performance in this context [6, 14].

Although trauma-specific models performed better overall, their predictive abilities were relatively close, suggesting substantial overlap in the physiological information they encode [6]. This indicates that current scoring systems may be capturing similar dimensions of injury severity rather than providing fundamentally distinct predictive insights. As a result, their clinical utility may depend more on practicality, speed of use, and institutional preference than on clear superiority of one system over another [5, 6].

The differences observed in diagnostic thresholds further emphasize the inherent trade-off between sensitivity and specificity across scoring systems. Models emphasizing broader physiological variables tend to identify a larger proportion of high-risk patients, while more neurologically focused approaches offer improved specificity. This balance is clinically relevant in emergency and intensive care settings, where decisions often involve prioritizing early identification of deterioration versus avoiding unnecessary escalation of care.

Despite these findings, the moderate overall performance of all evaluated scoring systems highlights an important limitation in current clinical prognostic tools [6, 9]. TBI is a highly heterogeneous condition influenced not only by initial physiological status but also by dynamic secondary injury mechanisms, intracranial pressure changes, imaging findings, and treatment-related factors [1, 7]. These dimensions are not fully captured by traditional scoring systems, which rely primarily on static admission variables [9].

In conclusion, early mortality in blunt traumatic brain injury is strongly associated with impaired consciousness and systemic physiological instability at presentation [1]. Trauma-specific scoring systems demonstrate better prognostic performance than general emergency department scores; however, no single model provides clearly superior or complete predictive accuracy. These findings highlight both the clinical utility and inherent limitations of existing scoring systems, which primarily rely on static clinical variables and may not fully capture the dynamic and multifactorial nature of secondary brain injury [9].

Future improvements in risk stratification are likely to depend on the development of more comprehensive, multi-dimensional predictive models that integrate clinical parameters with radiological features [19, 21], laboratory biomarkers, and potentially molecular or physiological time-series data [3]. Such approaches, particularly when combined with advanced analytical methods such as machine learning, may offer more accurate and individualized prediction of outcomes in patients with traumatic brain injury [3, 4].

## Statements

### Data availability statement

The raw data supporting the conclusions of this article will be made available by the authors, without undue reservation.

### Ethics statement

The protocol for this study was reviewed and approved by the Institutional Review Board of Iran University of Medical Sciences. The studies were conducted in accordance with the local legislation and institutional requirements. Written informed consent for participation in this study was provided by the participants’ legal guardians/next of kin.

